# Life-long music and dance relationships inform impressions of music- and dance-based movement therapies in individuals with and without mild cognitive impairment

**DOI:** 10.1101/2024.05.09.24307114

**Authors:** Meghan E. Kazanski, Sahrudh Dharanendra, Michael C. Rosenberg, Danyang Chen, Emma Rose Brown, Laura Emmery, J. Lucas McKay, Trisha M. Kesar, Madeleine E. Hackney

**Affiliations:** Department of Medicine, Division of Geriatrics & Gerontology, Emory University School of Medicine, Atlanta, GA, USA; Department of Medicine, Emory University School of Medicine, Atlanta, GA, USA; Department of Biomedical Engineering, Emory University & Georgia Institute of Technology, Atlanta, GA, USA; Rollins School of Public Health, Emory University, Atlanta, GA, USA; College of Arts and Sciences, Emory University, Atlanta, GA, USA; Department of Music, Emory University College of Arts and Sciences, Atlanta, GA, USA; Department of Biomedical Informatics, Emory University School of Medicine, Atlanta, GA, USA; Department of Neurology, Emory University School of Medicine, Atlanta, Georgia, USA; Department of Rehabilitation Medicine, Emory University School of Medicine, Atlanta, GA, USA; Atlanta VA Center for Visual & Neurocognitive Rehabilitation, Atlanta, GA, USA; Birmingham/Atlanta VA Geriatric Research Education and Clinical Center, Atlanta, GA, USA

**Keywords:** Alzheimer’s disease, mild cognitive impairment, rehabilitation, motor cognition, music therapy, dance therapy

## Abstract

**Background:** No effective therapies exist to prevent degeneration from Mild Cognitive Impairment (MCI) to Alzheimer’s disease. Therapies integrating music and/or dance are promising as effective, non-pharmacological options to mitigate cognitive decline.

**Objective:** To deepen our understanding of individuals’ relationships (i.e., histories, experiences and attitudes) with music and dance that are not often incorporated into music- and dance-based therapeutic design, yet may affect therapeutic outcomes.

**Methods:** Eleven older adults with MCI and five of their care partners/spouses participated (4M/12F; Black: n=4, White: n=10, Hispanic/Latino: n=2; Age: 71.4±9.6). We conducted focus groups and administered questionnaires that captured aspects of participants’ music and dance relationships. We extracted emergent themes from four major topics, including: (1) experience and history, (2) enjoyment and preferences, (3) confidence and barriers, and (4) impressions of music and dance as therapeutic tools.

**Results:** Thematic analysis revealed participants’ positive impressions of music and dance as potential therapeutic tools, citing perceived neuropsychological, emotional, and physical benefits. Participants viewed music and dance as integral to their lives, histories, and identities within a culture, family, and/or community. Participants also identified lifelong engagement barriers that, in conjunction with negative feedback, instilled persistent low self-efficacy regarding dancing and active music engagement. Questionnaires verified individuals’ moderately-strong music and dance relationships, strongest in passive forms of music engagement (e.g., listening).

**Conclusions:** Our findings support that individuals’ music and dance relationships and the associated perceptions toward music and dance therapy may be valuable considerations in enhancing therapy efficacy, participant engagement and satisfaction for individuals with MCI.

## INTRODUCTION

Individuals with Mild Cognitive Impairment (MCI) present measured declines in cognitive function beyond those expected due to aging that introduce subtle functional deficits without precluding independence during activities of daily living [1, 2]. While no effective therapies currently exist to prevent MCI from progressing to Alzheimer’s Disease (AD) [3], movement therapies integrating music and dance offer an effective, non-pharmacological option to offset cognitive decline [4, 5]. Such therapies are especially promising, as they couple physical activity with high cognitive engagement to mitigate the declines in *motor-cognitive integration* that manifest as impaired daily functioning [6–10]. However, stronger methodological rigor is needed to effectively implement these therapies for every individual, thus likely requires additional therapeutic parameters [11]. To improve the systematic design of such music- and dance-driven therapeutics, we must first deepen our understanding of individual-specific factors that likely impact therapeutic outcomes that are not often overtly incorporated as parameters in therapeutic designs, yet may affect treatment outcomes. These factors include individuals’ lifelong interactions with music and dance activities, accultured preferences and attitudes, perceived confidence and barriers, and perceptions of music and dance as therapeutic tools for physical and cognitive health.

Clinical interest in adapting existing music and dance practices for use as rehabilitation tools is bolstered by neurophysiological evidence documenting their motor-cognitive benefits [12–16]. Music is of clinical interest for its accessibility and versatility, spectra of engagement that ranges from active (e.g., composition, singing, playing an instrument) to passive (e.g., listening) [17], and its diverse positive benefits that include mood enhancement and reduced depression [18], enhanced episodic memory [19], improved verbal fluency and encoding [20], and elevated general cognitive function [21]. Dance offers cognitive stimulation coupled with motor complexity in the form of challenging, novel, and diverse multi-step movement sequences [22], with demonstrated benefits including increased neuroplasticity [23] and improved motor and cognitive outcomes critical for functional daily activities (e.g., balance, mobility, global cognition, and episodic memory) [24–31]. As dance intrinsically fosters enjoyment through social and creative expression [32], individuals with MCI engaging in dance-based therapies report alleviated anxiety [33] and depressive symptoms [32], as well as improved emotional wellbeing [34], evaluations of daily living [26] and quality of life [35].

Therapeutic convention is to adapt music- and dance-driven interventions to accommodate individuals’ or group’s abilities or levels of disease progression. In music-focused activities, including both active and passive modalities, therapeutic accommodations include musical instrument selection [36], adjusted musical complexity (e.g., rhythm, repetitiveness) [37], sensory considerations (e.g., volume, tempo, timbre) [38], and extent of physical movement incorporation [39]. Therapies incorporating music and dance often prescribe participants’ practice of precise, rhythmic movements based on an existing, codified dance form to conventional musical selections accompanying that form [31]. Curricula are extracted and modified from a broad variety of dance traditions including individual (e.g., aerobic, contemporary, folk) and partnered dance styles (e.g., Latin, Ballroom, Tango) [26, 29, 33, 40–47]. Therapeutic accommodations include incorporated safety measures, reduced intensity, modulated extent of motor-cognitive challenge (e.g., coordinative demands, movement complexity), and simplified meter and reduced tempo of musical selections [26, 40, 47–49]. Although options for adjusting therapeutic parameters to the individual or group are many, selecting the best accommodations remains a subjective process.

While studies reporting benefits afforded by dance- and music therapies may specify systematic prescription of adapted dance steps and broad accompanying musical selections parameters [25, 27, 50], studies rarely parameterize therapeutic administration to participants’ music and dance experiences and preferences [35, 49, 51]. For instance, music selections accompanying dance therapies are persistently underspecified: either selected at random from an instructor’s music library [35], only generally specified by type (e.g., “popular”, “relaxing”) [29, 33] or genre [47], specified in meter [26] or beats-per-minute, only [29], or reported with no information regarding accompanying musical selections, whatsoever [25, 27, 50]. While music and dance-driven therapies have shown promising neurological, motor-cognitive, psychosocial and behavioral benefits for individuals with MCI, this is despite an apparent lack of systematic tuning of therapeutic parameters to the music and dance-related preferences and experiences of participants.

Personal preferences in music genre or dance style may influence therapeutic responses to music- and dance-based interventions through emotional engagement, long-term memory cueing, and racial/ethnic differences in active engagement, and perceived capability. Music is deeply personal, being inextricably connected to individuals’ significant life events and places and stimulating physiological arousal, a sense of physical enjoyment and ability to communicate, and self-reflection when listening [52, 53]. This influence manifests in some music therapy contexts where music is actually selected in accordance with individual preferences and familiarity, serving to enhance emotional engagement and elevate dissociation to reduce pain [54, 55], promote neural activation patterns beneficial for emotional regulation [56], and alleviate agitation and depression in individuals with dementia [57, 58]. This phenomenon is in in accordance with Gerdner’s theory of individualized music intervention for agitation (IMIA: i.e., that personalized music reduces agitation events in individuals with AD better than classical, non-personalized music) [59]. By contrast, activation patterns undesirable for emotional regulation arise when introducing complexity, dissonance, and unexpected events in musical selections [56]. Further, musical passages relevant to the individual have the potential to cue long-term memory of movement sequences (e.g., dance steps from youth) [60]. Preferences for dancing or other forms of active music engagement has been evidenced as race/ethnicity specific [61, 62]. More broadly, familiarity and perceived capability — informed by the extent of an individual’s prior music or dance engagement — affect participants’ engagement with specific music and dance forms as therapy [63]. Thus, therapeutic parameterization of preferences and familiarity in music and dance forms must be critically considered as that which is meaningful and connected to the history of the individual.

Through cultural and lived experiences, individuals develop diverse perceptions, attitudes, histories, and preferences towards music and dance that constitute their “relationship” with these art forms. Features of these relationships may indeed be crucial considerations that may be incorporated to elevate therapeutic efficacy and engagement. Prior experiences with music and dance predict participant initial and continued engagement in art and music-based therapies in people with mental health disorders [63, 64]. Similarly, individuals with Parkinson’s disease who self-identified as dancers directly attributed their prior dance experiences toward a deep sense of meaning felt when attending dance therapy classes [65]. Thus, understanding individuals’ prior experiences and perceptions toward music and dance may be valuable in promoting therapeutic retention and engagement.

Outcomes of music- and dance-based therapeutics may be influenced by a participant’s “expectancy,” or personal belief that the therapy will elicit its intended benefits. Mechanisms supporting long-term engagement in dance-based programs in older adults with various health conditions included positive impressions of the therapeutic benefits, and expectancy that the dance delivers a good feeling about oneself, camaraderie and connection with others [66, 67]. Participants report perceived benefits following dance intervention participation including the opportunity to learn new skills, rediscover aspects of their younger selves, and connect with peers [68]. Although, it is unclear whether participants expected these benefits prior to study completion. Participants have also asserted that their self-efficacy in the ability to control their health through dance interventions have contributed to their engagement and adherence [69], with perceived benefits including freedom and increased self-expression [70]. This mood enhancing effect may be further augmented in a group setting [66, 67, 71]. Participants further report perceived benefits following dance interventions including opportunity to learn new skills, rediscover aspects of their younger selves, and connection with peers [68], although it is unclear whether participants expected these benefits prior to study completion. Participants who took part in each of dance or music programs asserted that their belief in the ability to control their health through the dance program played a big role in their adherence to and participation in the dance program [69] and that freedom and choice were large parts of their derived benefit from the exercise [70]. This mood enhancing effect may be further augmented in a group setting [71].

In this study, our objective was to characterize the individual-specific music and dance “relationships” (i.e., their histories, experiences, and attitudes towards music and dance) and the accompanying perceptions regarding music and dance as therapeutic tools for individuals with MCI. We conducted focus groups comprised of older adults with MCI and their care partners and spouses, in which we probed four major topics conveying lifelong relationships with an impressions of music and dance: (1) experience and history, (2) enjoyment & preferences, (3) confidence and barriers, and (4) impressions of music and dance as therapeutic tools. We extracted emergent themes within each of these topics. Accompanying subjective questionnaires evaluated similar features of participants’ music and dance relationships at the individual-specific level.

We expected that participants would share meaningful and formative experiences in various forms of music and dance through their lifetimes, and that they would connect these experiences to their diverse preferences in genre, style, and circumstances of music and dance exposure. We also expected that participants would report insightful barriers to engagement including circumstances (e.g., finances, health status, lifestyle) and social pressure and feedback (e.g., from instructors, family, peers). We further expected that participants would express enthusiasm about the potential efficacy of music- and dance-based therapies, particularly for cognitive benefit. Insights gained here will highlight the richness and depth of the music- and dance-related experiences, attitudes and preferences of individuals with MCI. These themes may identify music or dance-salient parameters that should be considered in the curated design of maximally efficacious, engaging, and enjoyable music- and dance-based therapies.

## METHODS

The Institutional Review Board at Emory University (STUDY#:00003507) approved this study. All participants provided written, informed consent prior to study participation.

### Focus Group Participants

Sixteen older adults participated. Eleven participants were screened and confirmed for amnestic MCI status, as defined by the Alzheimer’s Disease Neuroimaging Initiative criteria [72]. The remaining five adults were spouses or non-spouse care partners (e.g., relatives). Participants self-identified as 4 male/12 female and were racially diverse (Table 1).

**Table 1:** Focus group demographics, N=16 participants.

|  | *N (%) Mean (SD)* |
| --- | --- |
| **Age (y)** | **71.4 (9.6)** |
| **Sex** |  |
| Male | **4** |
| Female | **12** |
| **Cognitive Status** |  |
| MCI | **11** |
| Care Partners/ Spouses | **5** |
| **Education (y)** | **15.8 (2.5)** |
| **Race** |  |
| Black/African-American | **4** |
| White/Caucasian | **10** |
| Not reported | **2** |
| **No. of conditions** | **3.4 (1.4)** |
| **No. of medications** | **3.9 (3.0)** |
| **Hospitalizations in the past 3 years** |  |
| Yes | **5** |
| No | **11** |
| **No. of falls in the previous year** | **0.4 (0.8)** |
| **Fear of falling** (scale of 1-7, 7 indicates high fear of falling) | **2.3 (1.8)** |
| **Quality of Life** (scale of 1-7, 7 indicates high quality of life) | **6.4 (0.8)** |
| **Marital Status** |  |
| Married/Partner | **14** |
| Separated/Divorced | **2** |

Participants were required to have at least six years of education, English language proficiency, and no recent hospitalizations, medications, or medical or physical conditions that would preclude participation or affect cognition (e.g., major depressive disorders).

All participants completed questionnaires that surveyed demographic information, health history, and health status (Table 1).

### Music and Dance Relationship Questionnaires

We developed a Music Relationship Questionnaire (MRQ) to quantify each individual’s “relationship” (e.g., their history, experiences, perceptions, and attitudes) to music (Table 2, left). The questionnaire included ten items on a 7-category Likert scale to probe histories, daily experiences and engagement with passive and active forms of music. Two open-ended and one multiple selection items further probed music engagement history and accultured preferences, respectively.

**Table 2:** Music Relationship Questionnaire (MRQ) and Dance Relationship Questionnaire (DRQ) items.

| Item | MRQ | DRQ | Scoring Scale** |
| --- | --- | --- | --- |
| 1 | Music is important in my life. | Dance is important in my life. | Strongly disagree = 1<br>Strongly agree = 7 |
| 2 | I listen to music in my typical day. | I dance in my typical day.* | Strongly disagree = 1<br>Strongly agree = 7 |
| 3 | I focus on what I am hearing while listening to music. | While listening to music, I often physically respond (ex: tapping, moving, clapping, nodding, snapping). | Strongly disagree = 1<br>Strongly agree = 7 |
| 4 | I focus on other things while music is playing. | I do not often move while music is playing. | Strongly agree = 1<br>Strongly disagree = 7 |
| 5 | I actively choose the music I listen to. | I actively choose to move when music is playing. | Strongly disagree = 1<br>Strongly agree = 7 |
| 6 | I usually listen to whatever music happens to be playing. | I usually dance to whatever music happens to be playing. | Strongly disagree = 1<br>Strongly agree = 7 |
| 7 | I play an instrument/sing. | I dance regularly. | Strongly disagree = 1<br>Strongly agree = 7 |
| 8 | I have played an instrument/sung for most of my life. | I have danced for most of my life. | Strongly disagree = 1<br>Strongly agree = 7 |
| 9 | I played an instrument/sang only as a child. | I danced only as a child. | Strongly disagree = 1<br>Strongly agree = 7 |
| 10 | In a typical day, I play an instrument/sing. | In a typical day, I dance.* | Strongly disagree = 1<br>Strongly agree = 7 |
| 11 | Do you create your own music (compose, improvise, DJ, etc.)? If so, tell us what you do? | Do you create your own dances (choreograph, improvise, teach, perform, etc.)? If so, tell us what you do? | N/A |
| 12 | What kind(s) of music do you listen to?<br>☐ Classical ☐ Jazz<br>☐ Rock ☐ Heavy metal<br>☐ Pop ☐ Country<br>☐ Hip-hop ☐ Folk<br>☐ Rap ☐ Blues<br>☐ R&B ☐ Electronic<br>☐ Christian/Gospel ☐ Other | What kind(s) of dance do you mostly do, if any?<br>☐ Ballet ☐ Tap<br>☐ Ballroom ☐ Folk<br>☐ Contemporary ☐ Irish<br>☐ Hip-hop ☐ Modern<br>☐ Jazz ☐ Swing<br>☐ Other | N/A |
| 13 | Please describe an important musical experience and/or distinct memory connected to music and explain why. | Please describe an important dance experience and why it was important to you. | N/A |

\*Two DRQ items that were worded similarly and therefore were averaged before computing the composite DRQ score.
\*\*MRQ and DRQ scales were identical. Note that the Scoring Scale on Item 4 is reversed relative to all other items.

Similarly, we developed a Dance Relationship Questionnaire (DRQ) to quantify each individual’s “relationship” to dance (Table 2, right). The questionnaire was designed analogously to the MRQ, including ten Likert scale items probing histories, daily experiences and engagement with dance, as well as two open-ended and one multiple selection items probing dance engagement history and accultured preferences, respectively.

All participants completed the MRQ and DRQ prior to focus group participation. We quantified each participant’s “strength” of their music and dance relationships (1–7 scale) using composite scores, computed as the average of the ten Likert scale items of the MRQ and DRQ, respectively. Higher composite scores reflect stronger relationships.

### Procedures

We conducted two focus group sessions between July–August 2022. Each participant was sorted into one of two sessions based on their individual availability. Following informed consent procedures and survey/questionnaire administration, each focus group session was ∼90 minutes in duration.

Each participant was provided with a unique study ID (e.g., A1). Participants were referred to by their ID throughout the focus group session. Participants were identified with their ID throughout transcript analysis (Table 2).

The first focus group discussion included 6 participants, all provided with I.D.’s A##. The second focus group discussion included 10 participants, all provided with I.D.’s B##. Table 3 lists participant ID’s, stratified by demographics of race, biological sex, and MCI vs. Care Partner/Spouse status.

**Table 3:** Participant I.D.’s grouped by demographics of race, sex, and MCI vs. care partner/spouse status. I.D.’s beginning with A and B indicate participants of first and second focus group sessions, respectively.

|  | Female |  | Male |  |
| --- | --- | --- | --- | --- |
|  | MCI | Care Partner/<br>Spouse | MCI | Care Partner/<br>Spouse |
| **Black/African American** | A8, B1, B12 | - | B10 | - |
| **White/Caucasian** | A7, A9, B3 | A3, A4, B5, B7 | B4, B6, B14 | - |
| **Not Reported** | A5 | B13 | - | - |

One experienced focus group researcher moderated both focus group sessions. During the sessions, three study team members recorded field notes in real-time to capture any non-verbal interactions and noteworthy participant nonverbal behavior. Sessions were recorded and transcribed verbatim using Landmark (Landmark Associates, London, UK).

### Focus Group Guide

Focus group discussion topics were initiated using a focus group guide that consisted of a pre-determined series of question prompts. These prompts were designed to probe participants’ perceptions and experiences in areas including their music and dance: (1) “Experience & History,” (2) “Enjoyment & Preferences,” (3) “Confidence & Barriers to Engagement,” and (4) “Therapeutic Impressions” of music and dance-based interventions. Table 4 details the focus group guide and all prompts. The moderator strictly adhered to the prompted questions of the focus group guide throughout each session.

**Table 4:** Focus group guide with sample questions and prompts.

| **Key Topic:** | **Sub-topics:** | **Sample Probes:** |
| --- | --- | --- |
| **Experience and History** | **Role of Music:**<br>*What role has music played in your life?* | • **Music Genre Exposure:** *To what sort of music have you listened to or been exposed?*<br>• **Musicianship/ Instruction:** *Have you played musical instruments or sang? Were you formally instructed in music in any way?*<br>• **Music Experience:** *can you tell us an interesting or impactful musical experience?* |
|  | **Role of Dance:**<br>*What role has moving to music, or specifically dance, played?* | • **Dance Style Exposure:** *What dance or moving-to-music activities have you done? What structured moving-to-music or dance activities were you involved in throughout your life, if any? In what sort of movement AND music activities (e.g., gymnastics, dance, marching band) have you engaged?*<br>• **Dance Experience:** *Tell us an interesting dance experience.* |
|  | **Memories** | • *How do you think music can bring back memories for you? Can you describe an experience with that.* |
|  | **Music Inspiration** | • *When you hear the music do you feel motivated to move and do things?* |
| **Enjoyment and Preferences** | **Music Preferences** | • **Music Genre Preference:** *Tell me about some of your favorite types of music or songs.* |
|  | **Dance Preferences** | • **Dance Environment Preference:** *What environments do you like for dancing or moving-to-music?*<br>• **Dance Style Preference:** *What style of dance do you prefer?*<br>• **Dance Music Preference:** *What kind of music do you listen to when you dance?* |
| **Confidence and Barriers** | **Confidence Factors** | • *How comfortable are you with getting up and dancing in different situations (e.g. a bar, a party a fitness class)?* |
|  | **Music & Dance Challenges** | • *What, if anything, do you find challenging or intimidating about dancing or making music?* |
|  | **Feedback** | • *What sort of feedback did you receive from teachers, instructors, peers, or children about your dancing or music?*<br>• *Do you feel like the feedback that you have received from external sources has impacted your decision to continue to engage in music and dance?* |
| **Therapeutic Impressions** |  | • *How do you think music or moving to music could be used as a treatment for someone with memory impairments?* |

### Prescribed Topic Breakdown

All four major topics established in the focus group guide (Table 4) were discussed over 179 minutes of total discussion time. Figure 1 depicts the number of mentions for each prescribed major topic. The relative pattern of mentions across these prescribed topics demonstrate that the discussion focus generally reflected the number of questions administered in each prescribed topic, as outlined in Table 4. Of 563 total mentions, most of the focus group discussion was dedicated to participants’ music and dance “Experience & History” (276 mentions), followed by their “Enjoyment & Preferences” (150 mentions) then “Confidence & Barriers to Engagement” (94 mentions). A smaller proportion of the focus group was dedicated to discussions of participants’ “Therapeutic Impressions” of music and dance-based interventions (47 mentions).

**Figure 1:**
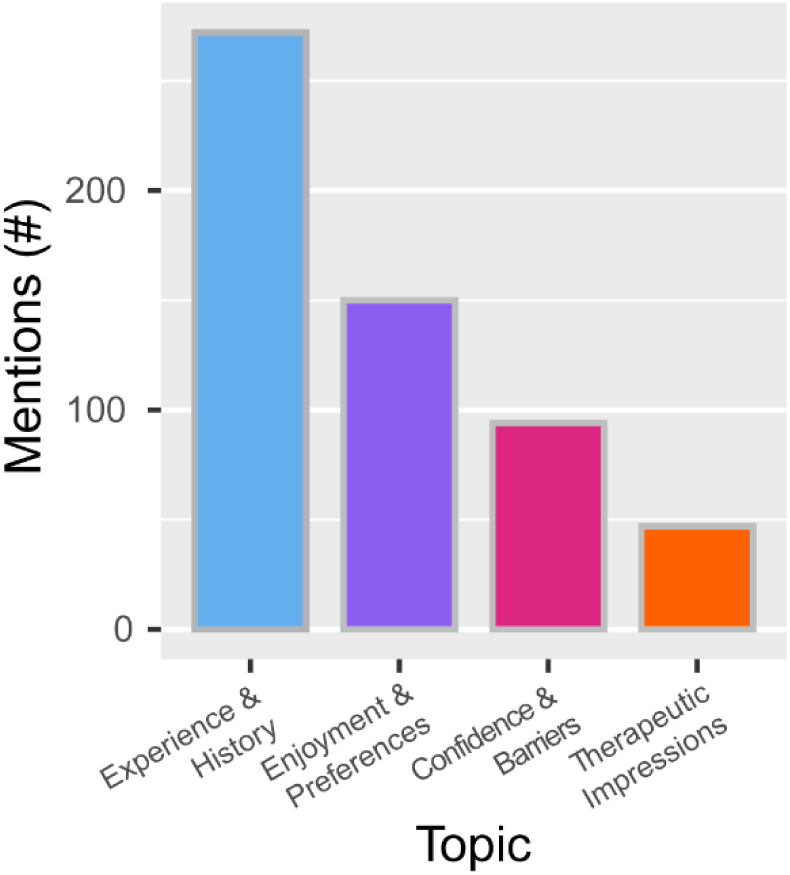
Overall distribution of mentions within each of 4 *a priori* defined topics of interest, as specified in the focus group prompt (Table 4). Frequency of mentions in each topic generally corresponds with the number of questions asked in that topic area. Thematic analysis was performed within each topic.

From each of the four prescribed topics, we extracted emergent major themes and nested sub-themes from the focus group transcripts. Discussion themes and sub-themes were defined by frequency-illustrative patterns, indicated by the number of times each theme and sub-theme occurred (i.e., mentions) in the discussion data. Within each topic, emergent themes and sub-themes were specified as any which exceeded a heuristically-prescribed 25 mention threshold, which accounted for at least 4% of the total discussion mentions.

### Transcript Analysis

Transcripts were analyzed in NVivo 12 software (Lumivero; Denver, CO). As in Lazris et al., [10], we applied a combined deductive and inductive analysis approach to extract themes and sub-themes from both focus group transcripts. We first organized the major topics pre-defined by the focus group guide (Table 4) into a deductive codebook. Two lab members independently coded transcript data into either these deductive nodes or into additional, inductively-defined nodes that captured emergent themes identified during the coding process. Any discrepancies between the two independently coded transcripts were reconciled by a third independent lab member not involved in the coding process. We quantified mentions of all participant responses coded into the final structure that included inductive and deductive nodes, which provided the basis for our final theme development. Themes and sub-theme mentions are visualized using histograms.

## RESULTS

### Participants

All participants (N=16) contributed to one of two focus group discussions. Participant demographics are summarized in Table 1. Eleven participants had an MCI diagnosis, while five were care partners or spouses to these individuals. All participants were well educated (15.8 ± 2.5 years), rated their quality of life as high (6.4 ± 0.8 out of 7 on the QoL-AD scale), and 14 of them were married. Participant I.D.’s are indicated in Table 2, stratified by sex and MCI diagnosis or care partner/spouse status.

### Music Relationship Questionnaire

Figures 2 illustrates participant subjective responses to the MRQ indicating each individual’s self-reported music relationship and music genre preference. Participant responses to the 10 questions in a 7-point Likert scale (Fig. 2A) indicate that most participants either somewhat agreed, agreed, or strongly agreed that music is important to their lives (87.5% participants), part of their typical day (75% participants), and something that they focus on when it is playing (75% participants). Further, most participants somewhat agreed, agreed or strongly agreed that they both actively choose the music they listen to (62.5% participants), but also listen to whatever music happens to be playing (75% participants). Most participants somewhat disagreed, disagreed or strongly disagreed that they have a play an instrument or sing (87.5% participants), have a history of doing so (75% participants), or do so in a typical day (75% participants). Participants’ composite scores across these 10 questions indicate they exhibited moderately strong relationships to music, with composite scores averaging 4.1 ±1.0 on a 1–7 scale, where 7 indicates the strongest music relationship (Fig. 2B). Participants responses to the MRQ multiple selection item indicate their accultured preferences for music genres primarily including Pop, Blues, Rock, and R&B (Fig. 2C).

**Figure 2:**
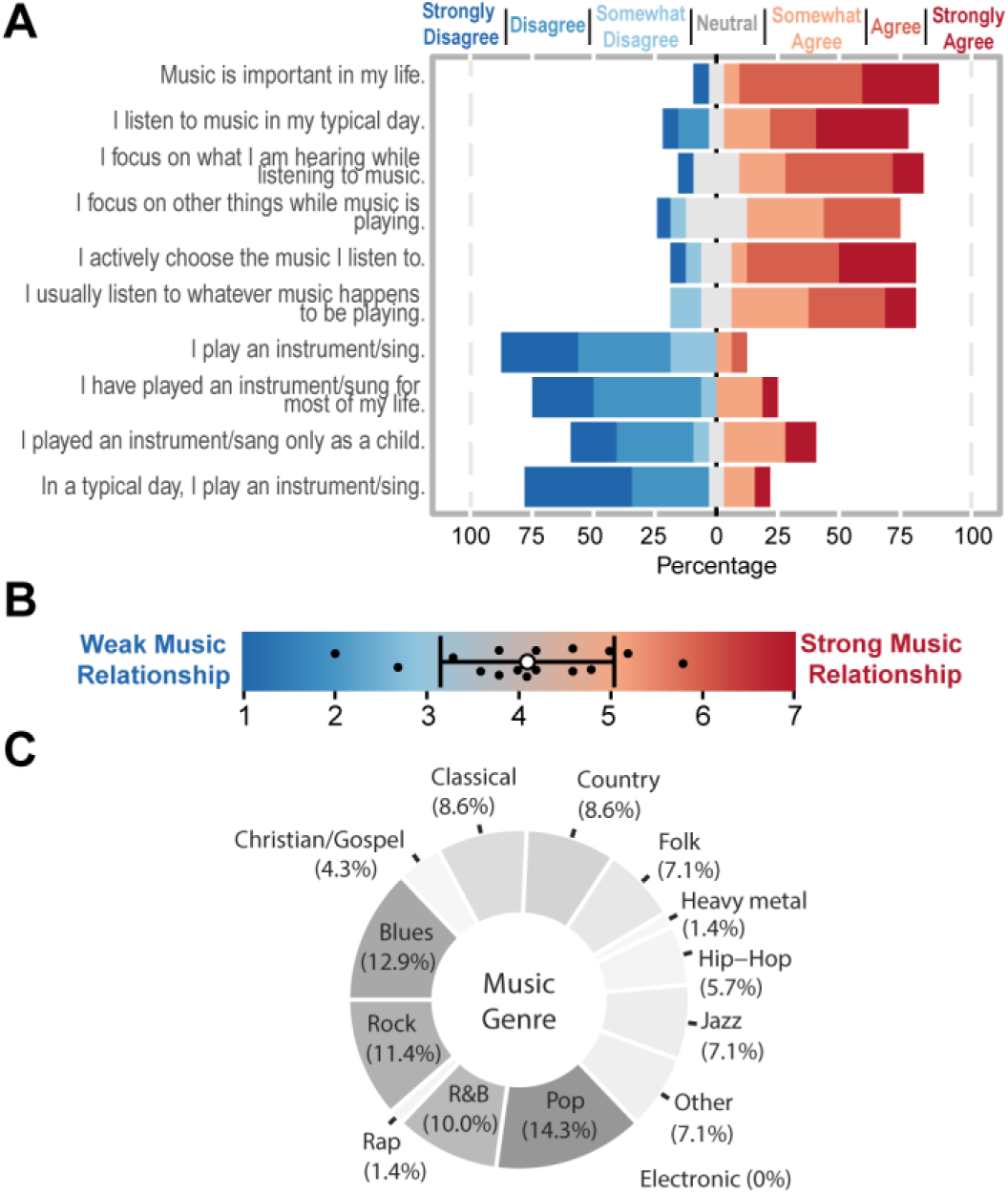
Participant responses to the Music Relationship Questionnaire (MRQ). (A): Divergence plots depicting group responses to each Likert Scale prompt on the MRQ, providing insight into their attitudes towards various aspects of music knowledge, including experience, proficiency, and usage in daily routine and task facilitation. Participants were generally in agreement that music was important to their lives and a part of their typical day, but tended to disagree with questions probing more active roles in music engagement (e.g., singing, instruments). (B): Bead-on-a-wire depicting individual (each black marker) composite scores across Likert Scale responses, representing the overall strength of their music relationships. White bead and black bars represent group mean and variance (±1SD) in composite MRQ scores. Composite score statistics indicate that participants, on average, had moderate relationships with music. (C): Donut plot representing group responses to preferred music genres indicated a preference toward Pop, R&B, Rock, and Blues.

### Dance Relationship Questionnaire

Figure 3 illustrates participant subjective responses to the DRQ indicating self-reported dance relationship and dance style preference. Participant responses to the 10-questions in a 7-point Likert scale (Fig. 3A) most notably indicate that most participants somewhat agreed, agreed or strongly agreed that dance is important to their lives (75% participants), and that they typically physically respond with movement when music is playing, e.g., clapping, tapping (100% participants). Participants either somewhat agreed, agreed, or strongly agreed that they actively choose to engage with music with this movement (87.5% participants) and will move to whatever music happens to be playing (75% participants). Participants either somewhat disagreed, disagreed or strongly disagreed that they do *not* physically respond when music is playing (62.5% participants) and that they danced only as a child (62.5% participants), although they do not dance in their typical day (62.5% participants). Participants’ composite scores across these 10 questions indicate they exhibited moderately strong relationships to dance, with composite scores averaging 4.6±0.8 on a 1– 7 scale, where 7 indicates the strongest dance relationship (Fig. 3B). Participants responses to the DRQ multiple selection item indicate their accultured preferences for dance styles primarily including Other, Contemporary, Ballroom, and Swing (Fig. 3C).

**Figure 3:**
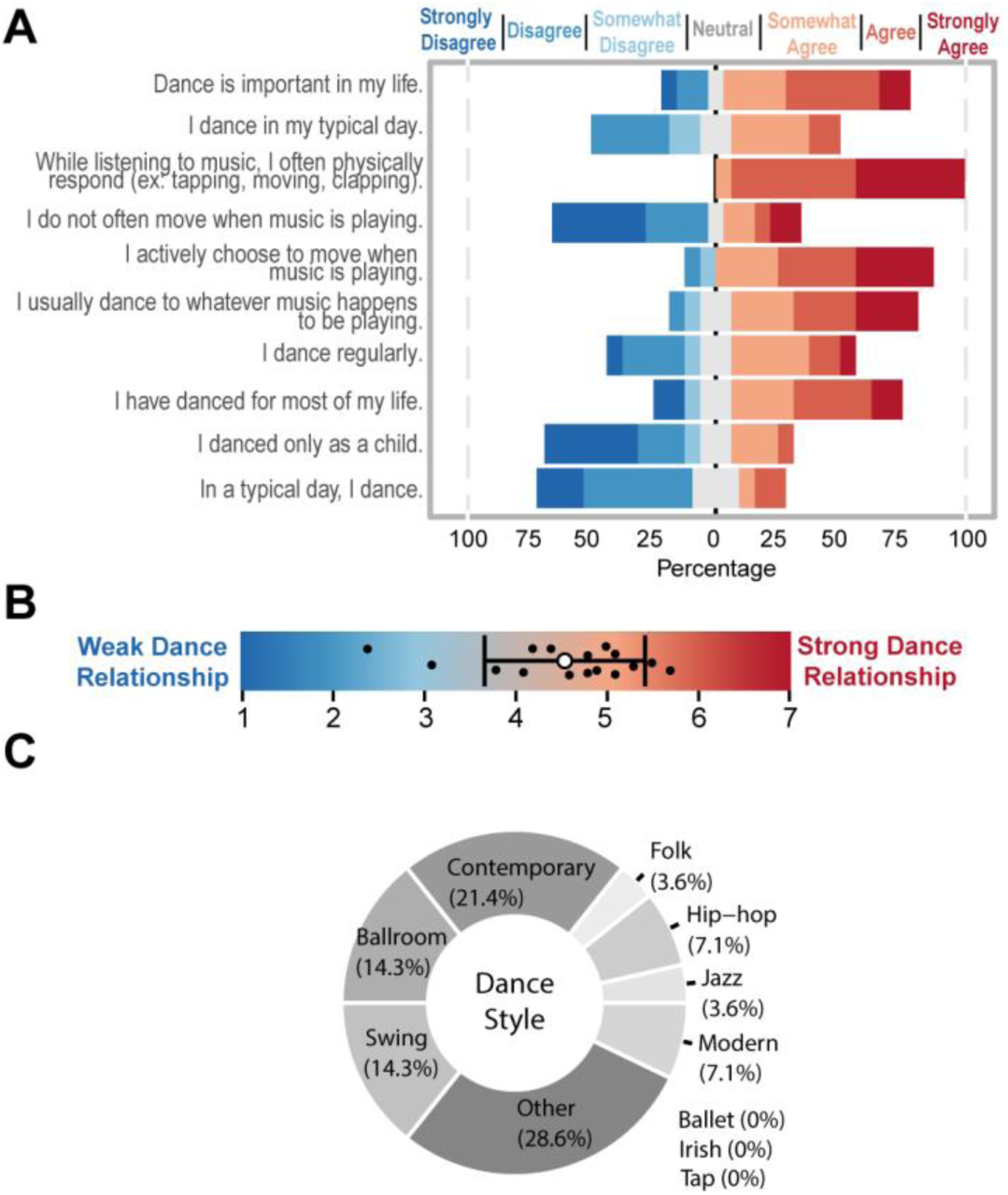
Participant responses to the Dance Relationship Questionnaire (DRQ). (A): Divergence plots depicting group responses to each Likert Scale prompt on the DRQ, providing insight into their attitudes towards various aspects of dance knowledge, including experience, proficiency, and usage in daily routine and task facilitation. Participants were generally in agreement that dance is important to their lives and that they often move to music that is playing, but tended to disagree that they only danced in childhood. (B): Bead-on-a-wire depicting individual (each black marker) composite scores across Likert Scale responses, representing the overall strength of their dance relationships. White bead and black bars represent group mean and variance (±1SD) in composite DRQ scores, respectively. Composite score statistics indicate that participants, on average, had moderate relationships with dance. (C): Donut plot representing group responses to preferred dance styles indicated a preference toward Contemporary, Ballroom, Swing, and other styles.

#### Topic 1: Experience & History

The topic resulting in the most frequent mentions (272 of 563 total) probed individuals’ experience and history of engaging in music- and dance-based activities, with the overall discussion breakdown represented in Figure 4A. Emergent themes included “Role of Music” (110 mentions) and “Role of Dance” (80 mentions). Participants also discussed memories that music and dance evoked, and articulated examples of how music inspires them to complete daily tasks (e.g., cooking, cleaning, driving), although these were not identified as major themes within this topic.

**Figure 4:**
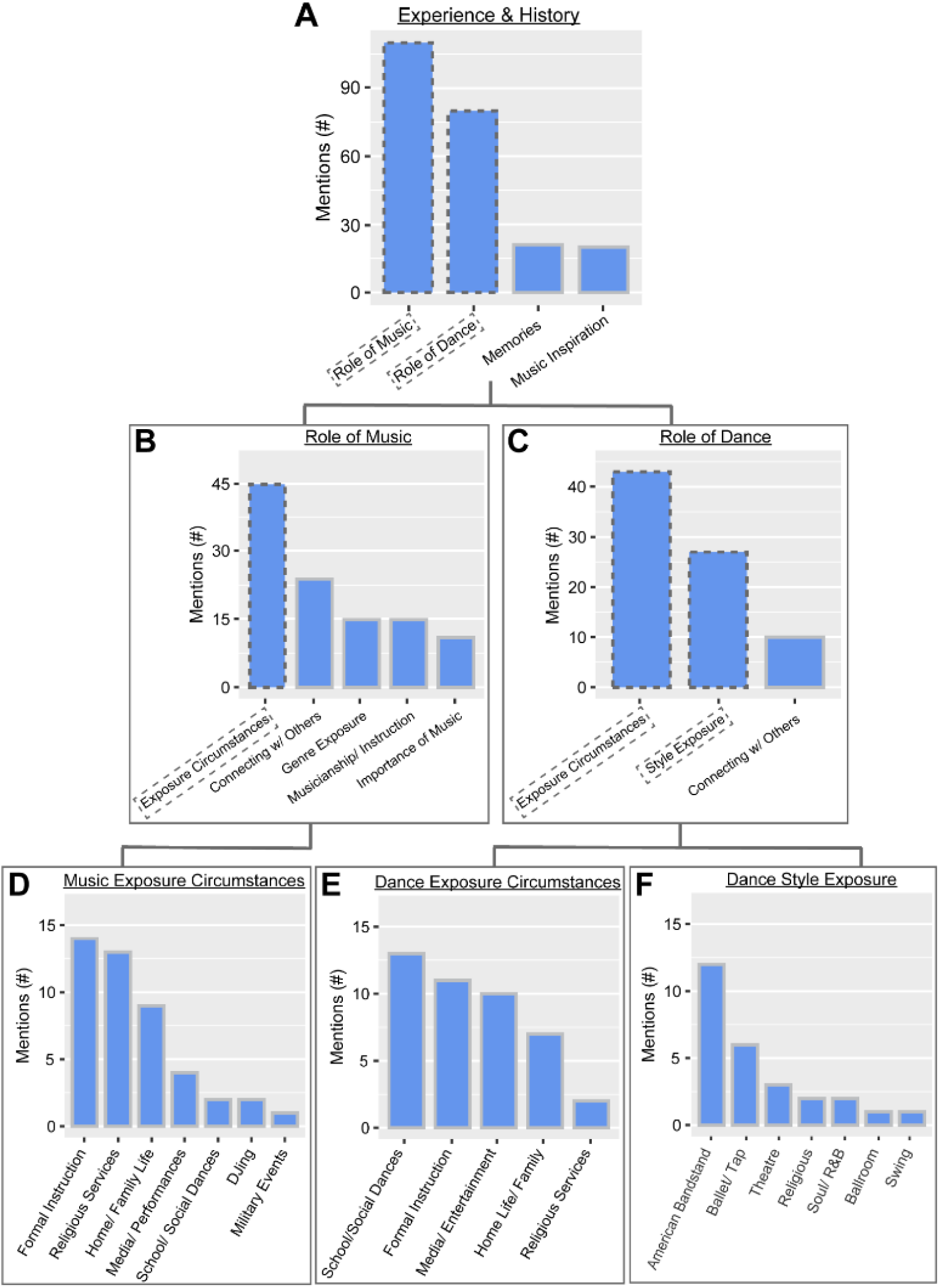
Distribution of mentions within prescribed Topic #1: “Experience & History” of music and dance. (A): Main themes pertaining to this topics included “Role of Music” and “Role of Dance,” but memories evoked, as ways that music inspires daily tasks were also recorded. (B): Distribution of mentioned within the “Role of Music” theme. One prominent sub-theme pertaining to this theme included “Exposure Circumstances,” while participants also mentioned how music connects them with others, specific genres to which they were exposed, musicianship and instrumentation experiences and the importance of music in their lives. (C): Distribution of mentioned within the “Role of Dance” theme. Prominent sub-theme pertaining to this theme included “Exposure Circumstances” and “Style Exposure,” while participants also mentioned how dance connects them with others. (D-F): Sub-themes were plotted to reflect distribution of mentions detailing participants’ experiences pertaining to their “Music Exposure Circumstances” (D), “Dance Exposure Circumstances” (E) and “Dance Style Exposure” (F).

### Theme 1-1: Role of Music

The primary emergent theme in the topic of “Experience & History” included “Role of Music” (Fig. 4B) in which participants offered specifics of the ways in which music has played a role in their lives. Given the large number of mentions in the “Role of Music” emergent theme (110 of 262 mentions), we classified “Music Exposure Circumstances” (45 mentions) as an emergent sub-theme for further elaboration. Mentions in this sub-theme detailed circumstances surrounding participants’ earliest and/or most impactful exposures to music. Participants also discussed topics pertaining to how music has played roles in their lives that did not emerge as sub-themes, including how music has connected them with others, their experiences in musicianship and instruction, and expressions of the importance of music.

### Sub-theme 1-1: Music Exposure Circumstances

Within the emergent sub-theme of “Music Exposure Circumstances” (Fig. 4D), participants most frequently referenced formal instruction as their means to music exposure (14 mentions). One participant detailed their brief bouts of music lessons from early childhood consisting of *“two years on the piano, and one on the clarinet,”* alluding to a stunted participation and enjoyment because of the associated challenge and detailing that these lessons *“did not progress into success ‘cause it was all work and no fun”* (A3). Another participant detailed the trajectory of their music exposure that conveyed their deep curiosity for music and desire to engage that began from a young age, then largely ended with low self-perceived efficacy as an adult. Speaking of music, this participant said:

*“It’s played an interesting role. When I was a little kid, I wanted to play the piano. I have no idea why. The lady who lived next door to us was gonna teach me piano, and then she had a heart attack the day before I was gonna start, so that ended that*.

*When I was in elementary school, I did—I sang in a choir. The older I got, I tried to do things that would get me into music. I did the choir, and I—one of my best friends played in the church, the organ, and I sat beside her and turned the pages. That got me a little closer to it. Then in high school, did all the typical high school thing. We had dances probably once every three weekends, and that was a great musical time for me*.

*Then when I was in college, I signed up for a piano class, learning—for people who had never taken piano, which was me. I walked in, and there were pianos all set up. The teacher walked into the room and sat down and said, “Hands on the keyboard. Ready? Play.” All of the maybe 15 people in the room started playing the music. I had no idea. That ended my instrument trials. [Laughter] I gave up”* (B5).

Other participants referenced religious services (13 mentions), including singing as part of church choir or congregation, as some of their earliest exposures to music. Throughout the “Music Exposure Circumstances” sub-theme, including in the two prior quotes, participants jointly detailed their earliest exposures to music with negative feedback experiences. As one participant recounts, *“I was in the children’s—in the boys’ choir at Independent Presbyterian church in Birmingham. When I hit puberty or shortly after hitting puberty, the choir director asked me just to mouth the words,”* (B4) to which the focus group moderator asked, *“Was that the end of your formal training?”* and the participant responded *“Yes. Though I do appreciate music”* (B4). Another participant mentioned that they joined the church choir in their youth after conveying their father’s misgivings about their participation in formal music instruction, recalling, *“My mom, who was determined for her five children to play the piano and sing…but my father finally put his foot down and said, “Okay, that’s it.” My brothers and I didn’t go any further with our education … I was in the church choir, but that was about it”* (B3).

Several participants noted a generational transmission of the roles of music in their earliest exposures to music in the home, from family (9 mentions). As one participant recalls exposure to music through family, *“My family, they are all musicians. They’re playing guitars; they’re singing; they write music. You know? There was music at home all the time. Singing and dancing and everything,”* then went on to detail how music was integrated with household tasks in their childhood and now in their current day-to-day, *“If I am cooking, I’m dancing… You’re sweeping with the rhythm of the music and singing. That’s the way we was brought up at home. Go to the river and do my laundry in the river; we didn’t have no washing machine… we’re just singing our heart out while we was doing our laundry”* (A5). Another participant recalled a similar ritual with music that has been transmitted across several generations, *“From a little girl, my father, every Saturday, we would get up and clean, and he would play music while we cleaned, so it was our ritual with that that I have passed over to my daughters”* (B13).

Participants also referenced circumstances including media, school and social dances, and military events that offered impactful exposure to music. Specific media referenced frequently included American Bandstand, with one participant citing, *“American Bandstand was my introduction to the popular music of the day. It was how I knew what to buy my next 45 record, or what the title of my next 45 record was gonna be ‘cause I bought one probably once a month. I saved up enough money to buy a 45 record. That’s how I knew what to buy”* (A7).

### Theme 1-2: Role of Dance

The secondary emergent theme in the topic of “Experience & History” included “Role of Dance” (Fig. 4C) in which participants offered specifics of the ways in which dance has played a role in their lives. Given the large number of mentions in the “Role of Dance” emergent theme (80 of 262 mentions), we classified emergent sub-themes including “Dance Exposure Circumstances” (43 mentions) where participants detailed the circumstances surrounding their earliest exposures to dance, and “Dance Style Exposure” (27 mentions), including participants’ earliest practiced and/or observed dance styles. Participants also discussed how dance has connected them with others, although this was not identified as a sub-theme.

### Sub-theme 1-2a: Dance Exposure Circumstances

Within the emergent sub-theme of “Dance Exposure Circumstances” (Fig. 4E), participants most frequently referenced school and/or social dances in their youths as their means to dance exposure (13 mentions). Participants especially noted feelings of belonging and enjoyment associated with these experiences, exemplified by one participant recalling, *“I started dancing, I think, when I was with other kids when I was in sixth or seventh grade, eighth grade, and then that was just rock and roll. It was like a club, but anybody could join. It was at a park or something. That was real fun, and I danced as long as I could”* (B6). Another participant expressed similar notions, *“All through high school, we also, after—on Friday nights, after whatever game it was, we always had to dance, and with live bands. There was this huge—and it really fostered a lot of dancing. I’m not sure I was that good, but I sure had fun”* (B5).

Additionally, participants prominently referenced formal dance instruction (11 mentions) as their means to dance exposure, but often also mentioned barriers associated with these early experiences. One participant referenced financial and familial challenges that disrupted her ability to participant in dance classes, *“My mother had me in ballet classes and everything. It didn’t last a whole bunch of time ‘cause she divorced and the money went. I continued with my daughter, with her dance classes and stuff”* (A9). Another participant mentioned a lack of interest, *“I really wasn’t interested in modern dance anyway. I had to take it—to fill in—plug in a course that I needed to have a class in it, and so I did it. I knew that I was not gonna pursue that any further”* (B5).

Participants also referenced media and entertainment (10 mentions), especially American Bandstand and Soul Train, in recounting their early dance exposure. For some participants, their concern was with keeping up with their peers in the latest dance styles, as one participant explains, *“Oh, American Bandstand used to come on 100 years ago. Then, of course, Soul Train took over. Do all my chores I got to do, get everything done, ‘cause I had to watch Soul Train to see who was on that. Then that’s when I learned the new dances ‘cause it’s, okay, what’s the new dance? ‘Cause I’m going to the club tomorrow, so I need to know how to do this dance, okay?”* (B12). Another participant mentioned American Bandstand as an outlet to access belonging amongst peers, *“I went to, you know, the popular American Bandstand and learned all of the dances and stuff…American Bandstand was the main place where I learned how to dance, learned all of the dances, and was able to—what I thought at the time—function like everybody else”* (A9). Other participants shared that American Bandstand *“did teach me a little bit about dancing”* (A8) and *“was important, and I did learn a lot”* (A4).

Participant mentions of early dance circumstances included in their home life (10 mentions), often while accompanying household chores, *“You’re mopping to the rhythm. You’re sweeping with the rhythm of the music and singing. That’s the way we was brought up at home”* (A5), or while bonding with family, *“I remember when we were growing up, how we would love—[mama] would put music on, and we would all dance because she loved to dance, so we loved to dance and listen to music and everything”* (B12). One participant recalled the foundational role of dance in their family life, recalling, “*Dancing played a big part in my life. My mother and my uncle used to be dance partners in dance contests. We had a lot of house parties when I was growing up, and everybody was dancing. Everybody, including my grandmother and grandfather. They would do the swing. It was just amazing*” (B1).

### Sub-theme 1-2b: Dance Style Exposure

Within the emergent sub-theme of “Dance Style Exposure” (Fig. 4F), participants most frequently referenced popular dance styles introduced to them through American Bandstand (12 mentions). Participants attributed their dance education and sense of belonging to this popular television program, as represented by one participant stating, *“Well, it’s how I learned to dance. The up-and-coming dances. You know? You’d grab a chair and you’d dance with it”* (A8). Another participant elaborated that American Bandstand was not only dance education, but a respite, elaborating, *“You’re right there watching it, so you can follow the steps much easier and learn ‘em quicker. I don’t know. I guess it was my relief from the woes and the goings-on around within the household”* (A9). Participants also mentioned exposure to ballet and tap, theatre, and dance at religious services. Notably, one participant recalls of some of the benefits and challenges that they encountered through exposure to clerical dancing in their church, detailing, *“I did learn a lot… I stayed in it until I got to the point where I could no longer remember the new steps. Dancing at church gave me some sense of having a better rhythmic movement. I’ve always enjoyed music. I love music. I’m just not very coordinated with it*” (A8). Other mentioned styles included Soul/R&B, Ballroom, and Swing.

#### Topic 2: Enjoyment & Preferences

The topic of “Enjoyment & Preferences” (150 of 563 total mentions) probed specific factors that contribute to participants’ enjoyment of music and dance experiences, including genres/styles, situations and environments, as well as general sentiments conveying enjoyment, apathy or displeasure toward music and dance experiences. The overall discussion breakdown is represented in Figure 5A. Emergent themes included preferred “Music Genre” (52 mentions) and “Sentiments” (45 mentions). Participants also discussed preferred environments and circumstances for engaging with music or dance, as well as preferred dance styles, although these were not identified as major themes within this topic.

**Figure 5:**
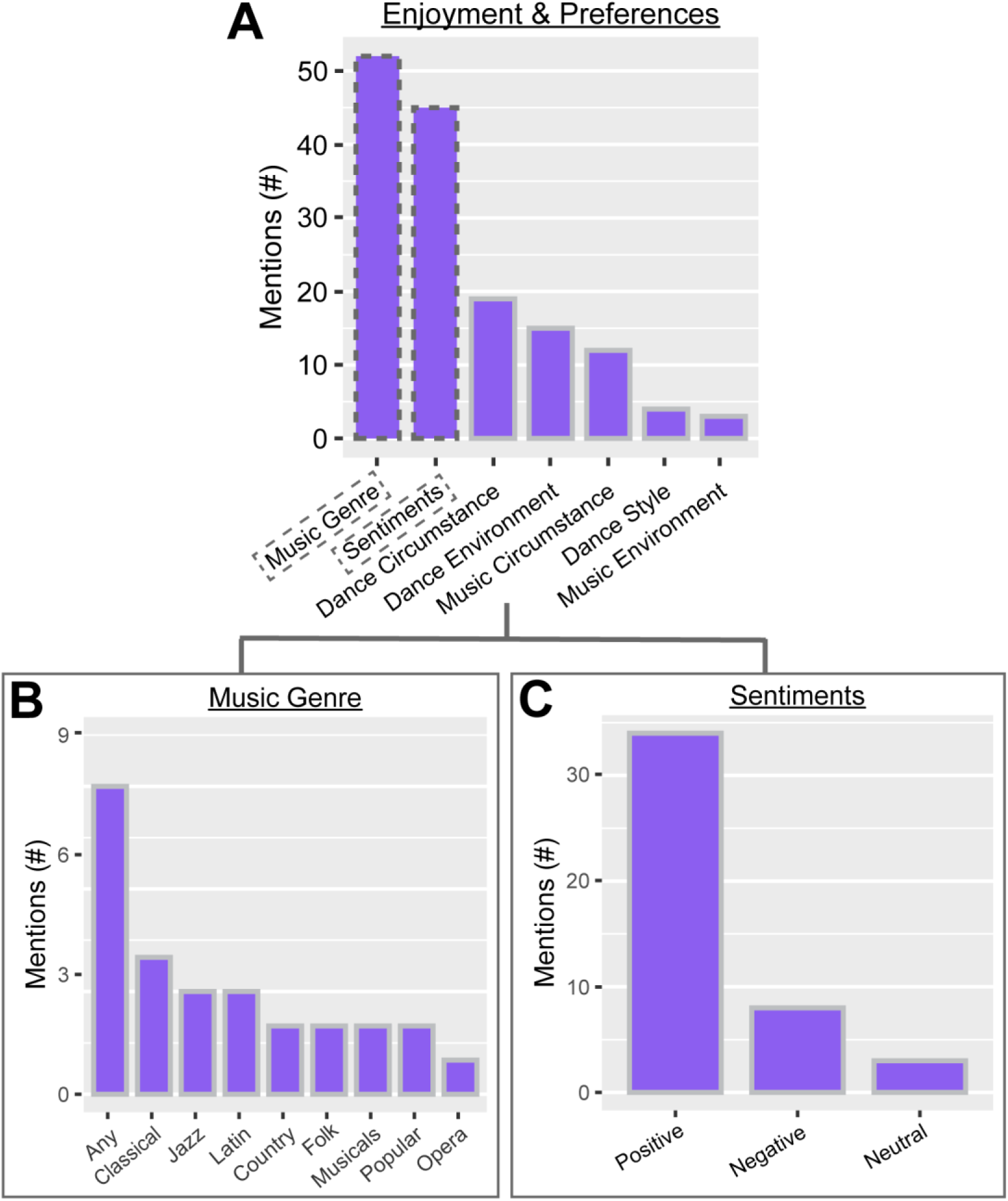
Distribution of mentions within prescribed Topic #2: “Enjoyment & Preferences” of music and dance. (A): Main themes pertaining to this topic including “Music Genre” and “Sentiments,” but specifics of preferred circumstances and environments in which to engage with music and dance, as well as preferred dance style were also discussed (B): “Music Genre” mentions reflect distribution of music genres preferred by participants, where several participants indicated no strong preference and reported Any genre. (C): “Sentiments” reflects mostly Positive, but some Negative, and few Neutral attitudes toward music and dance engagement.

### Theme 2-1: Music Genre Preference

The primary emergent theme in the topic of “Enjoyment & Preferences” included “Music Genre” (Fig. 5B) in which participants reported their music genre preference. Most responses indicated a general enjoyment of “Any” (9 responses) variety of music, with indications of slight preferences. For example, one participant stated “*I like all kinds of music. Everything. I loved to sing the music on the musical Fiddler on the Roof”* (A5). Another participant added, “*Again, I like all kinds of music. Everything. Any kind of music that was on at that time”* (A7), while referring to listening to the radio as a child. Responses across participants indicated engagement with different genres of music based on social settings such as family gatherings or religious groups.

Otherwise, participant preferences were varied across music genres including Classical, Jazz, Country, Folk, Musicals, Popular and Opera. One participant stated, “*I love Anita Baker, I like classical,”* (B1). Another participant mentioned, *“soul music, rhythm and blues”* (B12), while another added that, *“growing up I liked The Who, the Beatles, Rolling Stones”* (B4). One participant connected their music preference to their exposure to dance in their youth, stating, *“I learned to dance completely from American Bandstand. Any kind of music I really liked to hear, or to sing with or something like that”* (A9). Another specified genres that they did *not* enjoy, stating, *“I don’t really like hard rock, I’m not nuts about the electronic music”* (A4).

### Theme 2-2: Music & Dance Sentiments

Expressed sentiments toward music & dance were largely “Positive” (34 mentions), with some “Neutral” (8 mentions) and few “Negative” (3 mentions) responses (Fig. 5C). Responses conveying positive sentiments tended to reflect pleasant memories or social experiences. When asked about music, one participant stated, “*It just brings back happy or bad times, no matter which, depending on who you’re with and the song. That’s all it is. Usually it’s enjoyable whichever way it was because it’s a song”* (A4). Another participant added, “*Music is soothing and it helps the mind to relax, and then it brings back happy moments in your life”* (B1). When discussing how others in her life responded to her engagement with music, one participant noted that her child said, *“‘Mom, when you come home from choir practice, you are always so happy’,”* then went on to express, *“Music’s – just, it’s real important” (*B7).

#### Topic 3: Confidence & Barriers

“Confidence & Barriers” (94 of 563 total mentions) probed factors affecting individuals’ confidence in (e.g., perceived self-efficacy), as well as other barriers to engaging in music and dance-based activities. The overall discussion breakdown represented in Figure 6A. Emergent themes included “Barriers to Engagement” (45 mentions) and “Feedback” (41 mentions) and “Confidence Factors” (28 mentions). Participants also discussed challenges that they associated with music and dance, although these were not identified as major themes within this topic.

**Figure 6:**
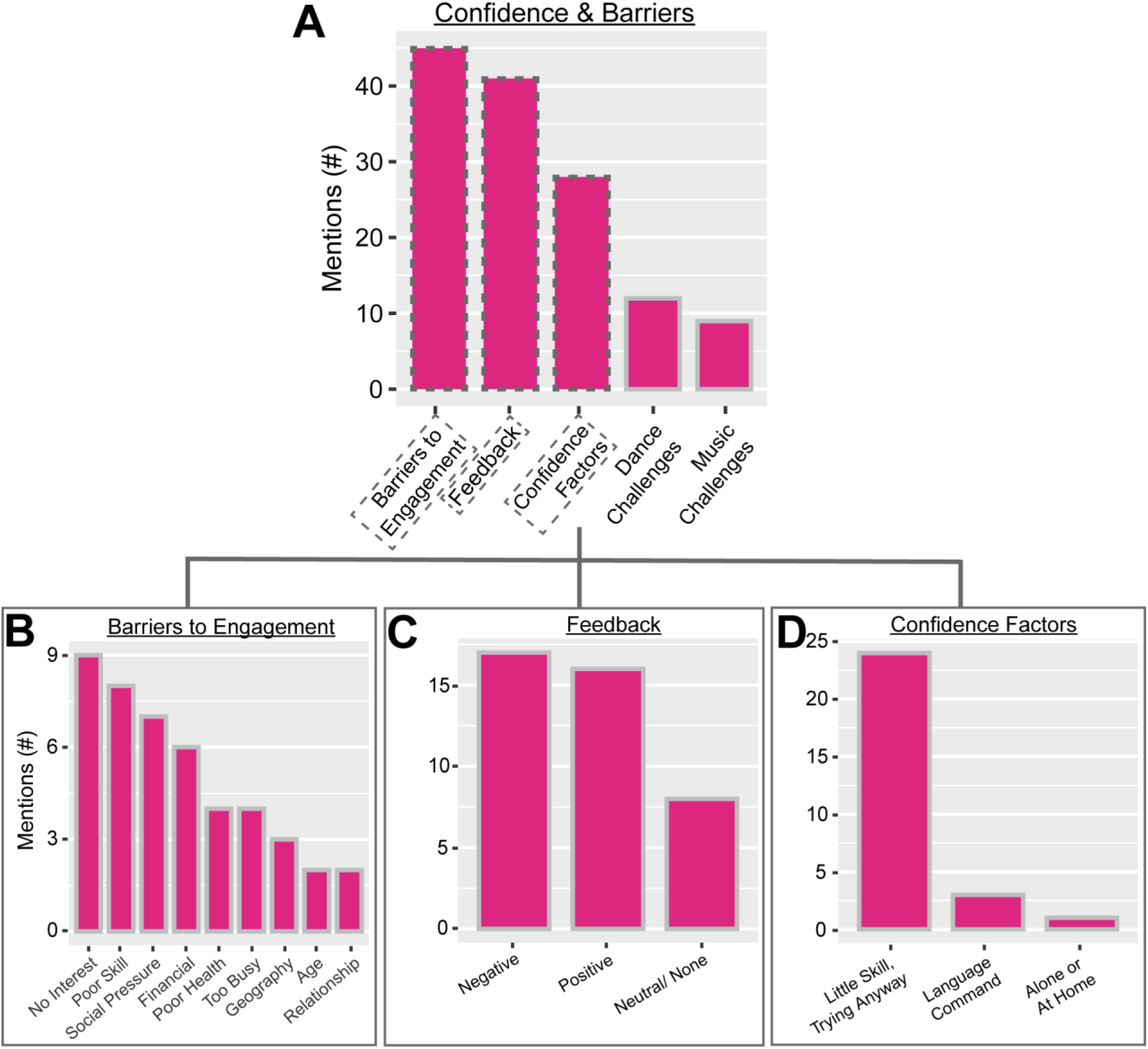
Distribution of mentions within prescribed Topic #3: “Confidence & Barriers” of music and dance. (A): Main themes pertaining to this topic included “Barriers to Engagement,” “Feedback” and “Confidence Factors,” while challenges associated with music and dance engagement were also discussed. (B): “Barriers to Engagement” mentions reflect distribution of identified factors limiting participants’ engagement with music and dance. (C): “Feedback” mentions reflect either “Negative,” “Positive” or “Neutral/None” feedback that participants received regarding the music and/or dance proficiency from family members, peers, or instructors. (D): “Confidence Factors” include factors impacting participants’ confidence while engaging with music and dance. Many participants noted a current stance of having little self-perceived ability, but having a willingness to engage, anyway.

### Theme 3-1: Barriers to Engagement

The primary emergent theme in the topic of “Confidence & Barriers” included “Barriers to Engagement” (Fig. 6B), in which participants shared specific factors that hindered their engagement with music and dance-related activities. Many individuals attributed their lack of engagement to “No Interest” (9 responses). This is exemplified by one participant explaining that their lack of engagement was largely due to a lack of perceived objective, stating, “*I don’t really see a goal in it; it’s not that I’m intimidated by it or anything ‘cause I can try to be ridiculous, but the aspect of it is I don’t get anything out of it*” (A3). This same participant continued, *“…some people wanna go play golf, some people wanna go bowling because they’re excited by it. I am just not excited by the music or the dance.”*

Other participants associated their lack of engagement with “Poor Skill” (8 mentions). Several participants recalled struggling to keep up with music courses or performance groups in their youth, then subsequently limiting their engagement because of either self-perceived and/or socially-instilled lack of proficiency. One participant recalled that their experience was limited by their perception of their musical proficiency in the first day of a college-level piano elective, with *“I walked in, and there were pianos all set up. The teacher walked into the room and sat down and said, “Hands on the keyboard. Ready? Play.” All of the maybe 15 people in the room started playing the music. I had no idea. That ended my instrument trials. I gave up”* (B5). Another participant recalled receiving pressure from a choir director to limit engagement due to lack of skill, *“When I hit puberty or shortly after hitting puberty, the choir director asked me just to mouth the words”* (B4). When the focus group facilitator asked whether that ended their formal training, this same participant responded, *“Yes. Though I do appreciate music.”*

Participants tended to also specify factors associated with early (e.g., childhood) barriers to engagement, including those arising from financial and geographical limitations, and social/familial pressures. For instance, participants recalled early barriers including that they *“never could afford a piano or any kind of instruments”* (B7), or having *“no facilities to go anywhere”* (A5), *“growing up in a third-world country”* (B10), or their family members *“laughing hysterically”* (B4) at them or a father *“putting his foot down”* (A3), thereby limiting music and dance engagement.

Other factors discussed were associated with later (e.g., adulthood, older age) barriers to engagement, including advancing age, too little time, and health problems. For instance, participants recalled more-recent barriers including *“old age”* (A8), being *“just so focused on working and making a living”* (A4), and multiple mentions of COVID-19. Another participant shared a personal health challenge, detailing, *“I can’t dance much anymore, but—because I have issues with my legs. They went bad, and I just—it’s not impossible, but it’s been tricky. It was very important for me”* (B6).

### Theme 3-2: Music & Dance Feedback

The secondary emergent theme in the topic of “Confidence & Barriers” included “Feedback” (Fig. 6C) in which participants detailed types of feedback that they received from family, instructors, and peers regarding their music and dance proficiency. Feedback mentions were most often “Negative” (17 mentions), with participants remarkably quoting specific feedback excerpts delivered to them from their dance and music instructors. As one participant recalls, their dance instructor once told them, *“‘this really isn’t your area’”* (B5). Other music instructors told participants that they, *“needed to stay quiet and weren’t carrying the tune”* (A4), or that they *“really don’t have feeling in the music”* (A3). One of these participants said that such feedback *“affected me a lot because, you know, then you think you’re no good”* (A4).

Participants also recalled impactful negative feedback from peers and relatives. Peers at school dances told one participant, *“‘You just have no rhythm whatsoever’”* and *“‘You really just can’t move’”* (A8). Another participant was told by their child in church, *“‘Shh. Mom, don’t sing’”* (A7). This participant cited this feedback as responsible for making them feel as though they are *“not allowed to sing in the congregation even in church.”*

Participants also recalled “Positive” feedback (16 mentions) from instructors, peers, and relatives. One participant recalls that a choir director encouraged them by saying, *“‘You have a beautiful voice, don’t worry about it’,”* (A7) which they later attributed to increased engagement and confidence in language learning, *“I learned music, and they start putting me to sing solos. Once I start singing the solo, it gave me more confidence in my language. You know? Here I am trying to learn more English.”* Several participants indirectly interpreted peers’ dancing with them as positive feedback regarding their dance proficiency, with one participant remarking, *“The only feedback that I can think of is that, okay, if somebody asked me to dance, it meant they knew that I could dance. That was the feedback”* (B5).

Other participants noted feedback classified as either “Neutral/None” (8 mentions), which was often accompanied by the notion of their peer-group’s consensus that proficiency was not important. One participant mentioned, *“I was just a part of a group, so I never did get any positive or negative feedback. Just they said, ‘We’re glad you’re here,’ kind of thing”* (B12). Another participant shared, *” Nobody was telling people, ‘You’re no good,’ or anything like that. It was just having fun. We weren’t doing it to learn to play music real—to dance real well”* (B5).

### Theme 3-3: Confidence Factors

The tertiary emergent theme in the topic of “Confidence & Barriers” included “Confidence Factors” (Fig. 6D) in which participants detailed specific factors that influenced their confidence while engaging with music and dance. Many participants noted low self-perceived efficacy, but an openness to try music and dance-related activities, anyway (24 mentions). As one participant explains, *“Even though I was not a good dancer and I never thought of myself as being a particularly good singer, it was something I enjoyed so I did it anyway”* (A8). Other participants echoed similar sentiments of engaging, despite perceived low-efficacy, stating, *“All through high school, we also, after—on Friday nights, after whatever game it was, we always had to dance, and with live bands. There was this huge—and it really fostered a lot of dancing. I’m not sure I was that good, but I sure had fun”* (B5).

Several participants explicitly mentioned that they did not find music and dance activities intimidating with an attributed fearlessness to trying new things, especially with increasing age. As one participant explained, *“I always would join a choir. I did it all even as I got older and everything. You know? I knew that I wasn’t sensational or anything like that, outstanding as a standout, but I could at least carry a tune. I did that, and the same with the dancing as I’m not afraid to attempt something new; and so that helped”* (A9). Another participant mentioned, *“In my early years, I was interested in getting the new steps, but now you just can just move slowly if you want to. As you get older, just keep up with the beat and do your own thing. No, I’m never intimidated about anything like that”* (B12). To a seldom extent, participants cited other factors, including “Language Command” and being “Alone or At Home” as things that increased their confidence in music and dance.

#### Topic 4: Therapeutic Impressions

The topic of “Therapeutic Impressions” was briefly discussed (47 of 563 total mentions) and probed individuals’ impressions of music and dance as therapeutic tools for individuals with memory impairments. The overall discussion breakdown of “Positive”, “Negative/Neutral” or “Negative” impressions is represented in Figure 7A. Emergent themes included “Positive Impressions” (44 mentions). Only 3 mentions were classified as either “Negative” or “Neutral/None”.

**Figure 7:**
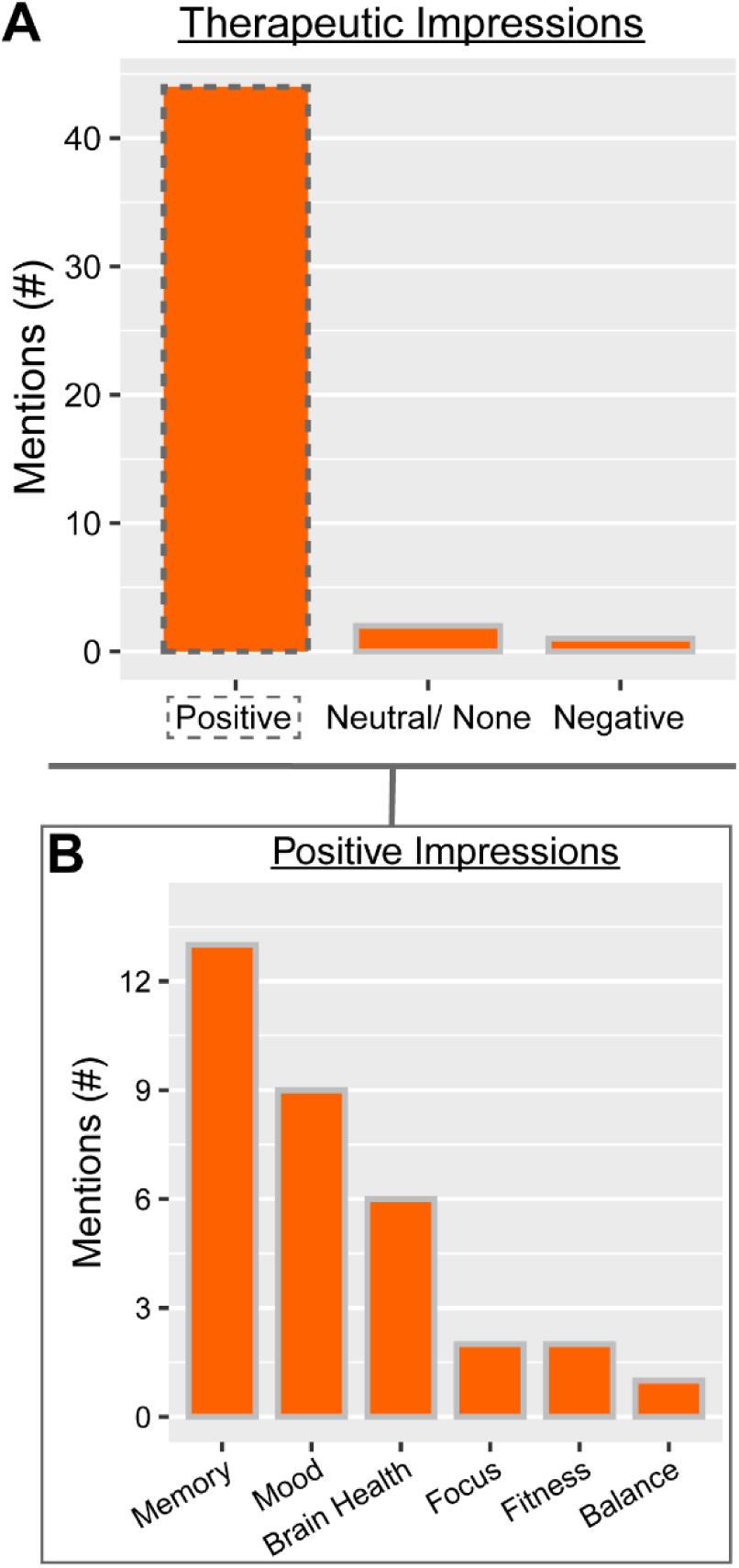
Distribution of mentions within prescribed Topic #4: “Therapeutic Impressions” of music and dance. (A): Main themes pertaining to this topic including “Positive Impressions,” but several neutral and negative impressions were also recorded. (B): “Positive Impressions” mentions reflect specific ways in which music and dance-based therapeutics could be beneficial.

### Theme 4-1: Positive Impressions

Within the emergent theme of “Positive Impressions” (Fig. 7B), participants discussed the ways in which music and dance-based therapies might benefit individuals with memory impairments. Participants most frequently expressed perceived benefits of music and dance on memory (13 mentions). Impressions of memory benefits are exemplified by one participant stating that music is “*a great way to bring things back in your memory, to exercise your memory, and that’s got to be good for you”* (B7). Another participant connected the benefits on memory to benefits on mood and brain health together, stating, “*Then on another level I think that people who have memory loss often have maybe depression and emotional problems. I think that any time you have music and light and communication with other people that it lifts you up, lifts your spirit, and again helps with your healthy brain function”* (A7). Another participant expressed their impressions of benefits of dance and music on brain health, and connected this to implications for memory, offering, *When you’re dancing, when you’re singing, you’re keeping your brain active. When you’re keeping your brain active, I’m thinking you won’t have a kind of memory loss because you are keeping moving”* (A5).

Participants further referenced perceived benefits on mood (9 mentions), with one participant stating that music is *“just so uplifting”* (B3), while another participant mentioned that music *“puts you in a happy space”* (B12). Another participant mentioned the uplifting and empowering capacity of music, expressing that the *“attitude”* of music is “*very upbeat”* and puts you in the headspace *“where you think, you can do just about anything”* (A9).

Participants expressed perceived benefits of music and dance on general brain health (9 mentions), with one participant recalling benefits that their younger sister, who was badly injured after a car accident, had following music therapy, stating, “*She came out of it when tapes were brought to her of music. It took a couple days, and she came out over it. I know that it has an effect on the brain, and I don’t know scientifically what that effect is, but I think it’s really good”* (B5). In this quote and throughout, participants were unclear on the exact mechanisms whereby music and dance positively impact health, but expressed belief in these benefits, nonetheless.

Other areas in which participants perceive positive impressions of music and dance included musculoskeletal health, focus, and balance. Notably, one participant elaborated on how they music helped them focus during their professional tasks, recalling, *“when I was a secretary for a group, and I had tons of reports that I had to get out every month. Without music, those reports would never happen…. As long as I had some music playing I could sit there and complete those reports and get them out on time”* (A8). Another participant shared his thoughts on how dancing might help slow the progression of his musculoskeletal disease, “*I think it would actually be good for my legs to get better maybe or not as bad as fast”* (B6).

Other participants espoused negative or neutral/no opinions regarding the potential for use of music and dance as therapy. One participant declared, *“it doesn’t help your muscles”* (A7), and another stated, *“I’m not sure about how it could be used for that,”* regarding the use of music and dance as therapeutic tools (A8).

## DISCUSSION

Our focus groups probed the lifelong music and dance relationships of older adults with MCI and their care partners and spouses. Using carefully designed prompts, we captured topics including participants’ music and dance: (1) “Experience and History,” (2) “Enjoyment & Preferences,” (3) “Confidence and Barriers,” and (4) “Therapeutic Impressions.” Participants expressed diverse, yet hopeful opinions about the potential for music and dance as therapeutic tools for individuals with memory complaints. One can surmise that these hopeful attitudes stem from largely positive music and dance relationships established over a lifetime of exposure and formative experiences. These participants saw music and dance as integral parts of their lives and histories that, despite being complicated by challenging experiences involving negative feedback or barriers to engagement, served to define them as part of a culture, family, and/or community.

The present study captured individuals’ limited engagement with active forms of music and dance that may be associated with their low self-efficacy in these areas. Both the focus group findings and MRQ and DRQ responses reflect participants’ enthusiasm for incorporating *passive* aspects of music and dance engagement into their lives with limited active engagement. These participants also detailed formative experiences that included negative feedback from family, peers or instructors. Participants indicated that this negative feedback often compelled them to disengage from and often develop low self-efficacy regarding their active music and dance activities. Self-efficacy factors are also broadly established determinants of treatment perseverance [73], engagement [74] and outcomes [75] in clinical practice. Our findings of low self-efficacy in these present participants suggest that in clinical practice, a given individual’ self-efficacy may be an important therapeutic consideration that could be incorporated into the design of music and dance-based therapies to maximize engagement and outcomes [77] and positively impact quality of life [78].

Our findings provide further support an individual’s impressions of therapeutic benefits or “expectancy” as a potential determinant of therapeutic process, satisfaction, and outcome [73, 75–78]. Participants in this present study reported overall positive impressions and high degree of expectancy in music and dance as therapeutic tools. These participants cited a variety of perceived benefits that ranged from cognitive factors (e.g., memory, brain health), to emotional factors (e.g., mood), to physical outcomes (e.g., musculoskeletal health, balance). These perceived benefits are aligned with desired rehabilitative outcomes identified by individuals with MCI and their caregivers [79, 80]. Characterization of the benefits recipients expect to receive, such as those described above, should be considered in therapeutic designs, as such perceived benefits may impact actual therapeutic outcomes [81]. Still, not all individuals with MCI and their care givers and spouses may view such therapies as potentially beneficial. For example, some may have greater expectancy or preference toward stage-dependent pharmacological or other established alternative approaches [82, 83]. However, as research evidence accrues supporting the benefits of music/dance therapies [84, 85], older adults with cognitive limitations may become increasingly receptive to music- and dance-driven therapeutics as primary or complementary approaches to supporting their cognition and general function.

The barriers to participation and negative feedback that participants sustained in their earliest music and dance experiences should be considered *alongside* their current openness and receptivity to engaging in music and dance activities. Notably, participant consensus largely identified a willingness to try engaging in music and dance despite a low perceived self-efficacy. This willingness presents therapist and clinicians with the opportunity to dissolve perceived barriers, and repair low self-efficacy associated with challenging earlier experiences by facilitating a positive, inclusive, and supportive environment for the music and dance-driven therapies. While specific parameters targeting improved self-efficacy through the treatment course are not yet defined, music and dance-based therapies offer improvements in psychological symptoms (e.g., anxiety, depression) [18] that are associated with self-efficacy [86] in individuals with AD [87]. Providing a warm, inviting setting founded on pillars of therapeutic alliance that include mutual respect, shared goals, and a strong focus on the participant’s treatment progress facilitates improved participant self-efficacy in other clinical contexts [88], and could be an effective approach in clinical music and dance therapies, as well. As evidence accrues supporting therapeutic efficacy and as national health organizations (e.g., the National Institutes of Health [89]) support the study of music and dance-based therapies, sensitivity to potential perceived and practical barriers to engagement is paramount.

The MRQ and DRQ quantitatively captured, at the *individual level*, similar notions as those qualitatively captured in the focus group discussions at the group level regarding the relative importance of individuals’ relationships with music and dance and their preferred forms of engagement. Participants designated music, especially in the passive domain, to be an important part of their lives. Participants expressed a similar regard for the importance of dance. During focus group discussions, participants shared impactful and sometimes fond memories of dance and other forms of active engagement with music (e.g., playing instruments, singing) from their youths, although mentions of recent or current participation in dance or other active forms of music engagement were lacking. Participants cited daily experiences including driving, cleaning, and focused work as examples in which passive engagement with ambient music elevates their *current* day-to-day lives. These notions were reflected at the quantitative individual level through MRQ and DRQ responses, where composite MRQ and DRQ scores both indicated moderately-strong relationships, on average, with each of music and dance. These results together highlight the ubiquitous presence of music and dance in individuals’ lives that tend to be more active and dance-inclusive in youth, then transitions to more passive, ambient music engagement into older age. This time-varying course of engagement with music and dance should be considered in the therapeutic integration of music and dance components, as individuals will likely be most receptive to those active forms of dance and music engagement with which they have some familiarity, likely from their youths. The administration of survey tools such as the MRQ and DRQ as we have demonstrated here, or similar [90, 91], critically discern the particulars of an individual’s current and past engagement and their genre and style preferences to succinctly offer valuable therapeutic design insights regarding the unique participant’s status, experience and preferences.

Participants’ decisive and varied preferences in music genres and dance styles, both articulated in focus group discussions and succinctly at the individual level in the MRQ and DRQ, may be harnessed to improve personalized therapy curation. Notably, participants’ preferences were often founded on pleasant memories and earlier experiences of music and dance. A recurrent example was an indicated preference for the popular music and dance styles that participants encountered on *American Bandstand* in their youths. Given the well-established benefit of familiar music on autobiographical recall in individuals with cognitive limitations [92–94], and the enhanced psychosocial and behavioral outcomes that arise from tuning therapeutic parameters to participant preference [95], these preferences identified herein and the accompanying prompts for extracting them offer valuable direction for explicit parametrization of participants’ preferences in music and dance-based therapies. Encouragingly, many participants indicated that they did not have a strong music preference and were open to many genres. This could be of immense benefit in group-class environments [22, 47, 96] where group consensus on preferences requires accommodation.

### Limitations

This study has several limitations. The sample size (N=16) is relatively small and is a heterogeneous mixture of mostly individuals with MCI, but also their care givers and spouses. This reduces our ability to make definitive conclusions regarding the perspectives of exclusively individuals with MCI who would be the recipients of music and dance-based interventions. The group that was assembled for this study had diverse backgrounds in engagement with music and dance therapeutically, as some participants had attended 20+ therapeutic dance sessions prior to study participation, while others had attended none Whether anyone had engaged in music therapeutically outside of a dance class is unknown. Further, the perspectives of this sample of individuals with MCI and their caregivers and spouses, while relatively diverse in biological sex and race and thereby likely from varied cultural backgrounds, is representative of the Atlanta metro area only. Given the role of culture in shaping exposure and responses to music and dance as culturally ubiquitous, yet diverse art forms [97, 98], a focus group held in another part of the US, or in another country may reveal different music and dance relationships from those we present here. We thus posit that this work should serve as a template for future explorations of this rich topic in other parts of the United States and the world. Further exploration will establish the generalizability of these present findings.

### Conclusions

This work highlights the richness of individuals’ *life-long* relationships with music and dance that may advise design of music- and dance-based therapies: in particular, how histories in music and dance starting from childhood contribute to individuals’ self-efficacy, present engagement, and expectations from therapies that incorporate music and dance. Participants’ engagement, preferences and attitudes may be investigated qualitatively via collective focus group responses or quantitatively via individual responses to objective music and dance relationship questionnaires, as both approaches afforded similar insights in the present study. We recommend that clinicians, in their own therapeutic design, consider the most impactful features of individuals’ music and dance experiences that emerged from these present discussions with individuals with MCI and their care partners/spouses, some of which included: barriers to engagement that permeated throughout their lifetimes, negative feedback from family, peers, and instructors that instilled persistent low self-efficacy, and how specific genres and styles align with individuals’ connections felt with loved ones, peers, and their community. Importantly, these features are accompanied by individuals’ openness to engaging in music and dance-based activities regardless of their perceived skill level, citing a variety of anticipated health benefits. Together, these factors serve to more-holistically characterize individuals’ relationships with and preferences in music and dance. We invite clinical implementation of music and dance-based interventions to consider this present work as a template to comprehensively probe participants’ relationships, through focus groups, MRQ and DRQ administration, or both. These insights may be incorporated as therapeutic considerations to likely enhance therapeutic efficacy, engagement and enjoyment in music and dance-based therapies targeting individuals with early cognitive decline.

### CRediT Author contributions

**Meghan E. Kazanski** (Data Curation; Methodology; Formal Analysis; Software; Validation; Visualization; Writing – original draft; Writing – review & editing)

**Sahrudh Dharanendra** (Investigation; Formal Analysis; Software; Writing – original draft; Writing – review & editing)

**Michael C. Rosenberg** (Investigation; Visualization; Writing – review & editing) **Danyang Chen** (Software; Formal Analysis; Visualization; Writing – review & editing) **Emma Rose Brown** (Software; Formal Analysis; Writing – review & editing)

**J. Lucas McKay** (Conceptualization; Data Curation; Funding Acquisition; Methodology; Supervision; Writing – review & editing)

**Laura Emmery** (Conceptualization; Funding Acquisition; Methodology; Supervision; Writing – review & editing)

**Trisha M. Kesar** (Conceptualization; Funding Acquisition; Methodology; Supervision; Writing – review & editing)

**Madeleine E. Hackney** (Conceptualization; Data Curation; Investigation; Funding Acquisition; Methodology; Validation; Project administration; Resources; Supervision; Writing – review & editing)

## Data Availability

All data produced in the present study are available upon reasonable request to the authors.

## Acknowledgements

We thank all volunteers who participated in the study, as well as T. Prusin, C. Carroll-Sauer and G. Harris for their assistance in data collection & participant recruitment.

## Data availability

The data supporting the findings are available upon reasonable request.

